# Sonication of the Anterior Thalamus with MRI-Guided Low Intensity Focused Ultrasound Pulsation (LIFUP) Changes Pain Thresholds in Healthy Adults: A Double-Blind, Concurrent LIFUP/MRI Study

**DOI:** 10.1101/2020.04.08.20042853

**Authors:** Bashar W. Badran, Kevin Caulfield, Sasha Stomberg-Firestein, Philip Summers, Logan T. Dowdle, Matt Savoca, Xingbao Li, Christopher W. Austelle, E. Baron Short, Jeffrey J. Borckardt, Norman Spivak, Alexander Bystritsky, Mark S. George

## Abstract

**Background:** Low Intensity Focused Ultrasound Pulsation (LIFUP) is a noninvasive brain stimulation method that may modulate deep brain structures using ultrasonic waves. Presently there are limited studies in humans rigorously assessing behavioral effects following LIFUP sonication of deep brain nuclei. As an initial test, we investigated whether sonication of the anterior thalamus, a central relay structure of nociception, would modulate thermal pain thresholds in healthy individuals.

**Methods:** We enrolled 19 healthy individuals in this three-visit, double-blind, randomized, sham-controlled, crossover trial. Participants attended a first MRI screening visit to acquire anatomical scans for LIFUP targeting. They then attended two identical experimental LIFUP/MRI visits (counterbalanced by condition) at least one-week apart. Within the MRI scanner, participants received two, 10-minute sessions of either active or sham LIFUP spread 10 minutes apart to the right anterior thalamus [Fundamental frequency:650khz, pulse repetition frequency: 10 HZ, Pulse Width: 5ms, Duty Cycle: 5%, Sonication Duration: 30s, Inter-Sonication Interval: 30 s, Number of Sonications: 10, ISPTA.3 719 mW/cm^2^]. Each 10-minute session was delivered in a block design (30s ON, 30s OFF). The primary outcome measure was quantitative sensory thresholding (QST), measuring sensory, pain, and tolerance thresholds to a thermal stimulus applied to the left forearm before and after LIFUP. Thermal stimuli were also applied in the scanner during certain blocks, either alone, or during LIFUP.

**Results:** Two 10-minute sessions of thalamic LIFUP produced a significant antinociceptive effect on pain thresholds. Temperature sensitivity increases were significantly attenuated (timeXcondition p=0.046) after active LIFUP (0.51 degree change) relative to sham stimulation (1.08 degrees). LIFUP also changed sensory and tolerance thresholds mathematically but this was not statistically significant with this sample. LIFUP delivered concurrently with thermal pain had no immediate behavioral effect.

**Conclusions:** Two, 10-minute sessions of anterior thalamic LIFUP has antinociceptive effects in healthy individuals. Future studies should optimize the parameter space and dose and perhaps investigate multi-session LIFUP interventions for pain disorders.

## Introduction

Although the field of brain stimulation has many different methods and approaches to modulate the central nervous system, newer techniques are needed that improve on the current toolkit. Transcranial magnetic stimulation (TMS) is FDA-approved for treating major depression and OCD, but can only focally stimulate the superficial cortex. While increasing the intensity of TMS or using broader deeper coils^1^ allows for greater depth of stimulation, this has the consequence of reducing focality.^2^ In contrast, deep brain stimulation (DBS), which is FDA-approved for treating Parkinson’s Disease, can focally stimulate deep in the brain.^3^However DBS is invasive and requires brain surgery. One promising new technology that is non-invasive and may be focally applied even deep in the brain is low-intensity focused ultrasound pulsations (LIFUP).^4,5^ LIFUP uses transducers containing piezoelectric elements that produce pulses of ultrasonic waves that summate deep in the brain.^6,7^ LIFUP is particularly intriguing as it is a noninvasive way of stimulating deep in the brain in a relatively focal area, a combination that is difficult to achieve with other technologies and could have substantial research and clinical potential for LIFUP.^4,5,8^

Prior preclinical research has helped to establish LIFUP safety limits and suggests that LIFUP is a safe and effective means of neuromodulation ^9-13^. Importantly, previous studies using stimulation doses similar to the current study have not observed characteristic negative physiological changes within the sonicated area such as cavitation or heat damage ^14^. Researchers are also beginning to use LIFUP in humans, studying how different stimulation parameters can induce reversible physiological effects on the nervous system, ranging from increased excitation in regions of interest ^15^ to suppression of visual evoked potentials ^16^.

We have chosen to investigate whether LIFUP can modulate pain as society needs non-opioid methods of modulating pain. While safe and successful in most clinical settings, opioid anesthetics are limited by dose-dependent side effects and potential abuse ^19 20^. U.S. emergency room visits and fatal poisonings caused by nonmedical use of opioid analgesics have more than doubled and tripled, respectively, within the last decade ^21,22^. Extrapolating off of the existing body of research, we wanted to investigate if LIFUP could modulate the anterior nuclei of the thalamus that are involved in pain perception.^17,18^ If LIFUP delivered to the anterior thalamus caused a measurable change in pain perception, it would help to validate LIFUP as a neuromodulatory device in humans, and serve as a stepping stone to a potential acute or chronic pain therapy. As a first step, we investigated whether MRI-guided sonication of the anterior thalamus could alter pain perception in healthy adults and whether LIFUP could produce quantifiable behavioral effects from a deep brain target.

Using a validated thermal pain paradigm within the fMRI scanner, ^23-26^ we hypothesized that LIFUP would be antinociceptive via modulation of the anterior thalamus. In this paradigm, anti-pain interventions require subjects to need a hotter temperature to feel the sensation, or label it as pain, or press a button to stop the stimulus.

## Methods

### Study Overview

We conducted a three-visit, double-blind, randomized, sham-controlled crossover trial in this concurrent LIFUP/MRI study. We recruited 29 healthy subjects between 18-45 years old from the local community through email broadcast. Exclusionary criteria included the following: seizure history (individual or family), history of depression, hospitalizations or surgeries in the previous 6 months, currently experiencing pain or a history of chronic pain, pregnant, alcohol dependent or any illicit drug use in the previous 6 months.

During their initial visit, participants were screened and signed written informed consent. Of the 29 consented, 19 completed the study and yielded usable data (11 women, mean age = 24.5 SD 4.6, range =18-38). All procedures were reviewed and approved by the MUSC Institutional Review Board.

After screening and signing MUSC approved written consenta T1-weighted MPRAGE (TR: 2300ms TE: 2.32ms, TI: 900m, acq time: 5:21, 0.9mm isotropic) anatomical image was acquired for each participant. This MPRAGE scan was used in conjunction with Brainsight neuronavigation (Rogue Industries). Neuronavigation outside of the MRI scanner using this structural scan allowed for the determination of the optimal location on the participant’s scalp that would position the LIFUP transducer to direct a sonication beam to the right anterior thalamus, considering both beam angle and depth.

Participants subsequently returned on two separate experimental LIFUP/MRI visits at least one week apart during which they received two, 10-minute sessions of either active or sham LIFUP within the bore of the MRI scanner (**Figure 1**). Participants were randomly assigned to receive either active or sham LIFUP the first visit and the opposite the second visit in a counterbalanced design.

**Figure 1.**
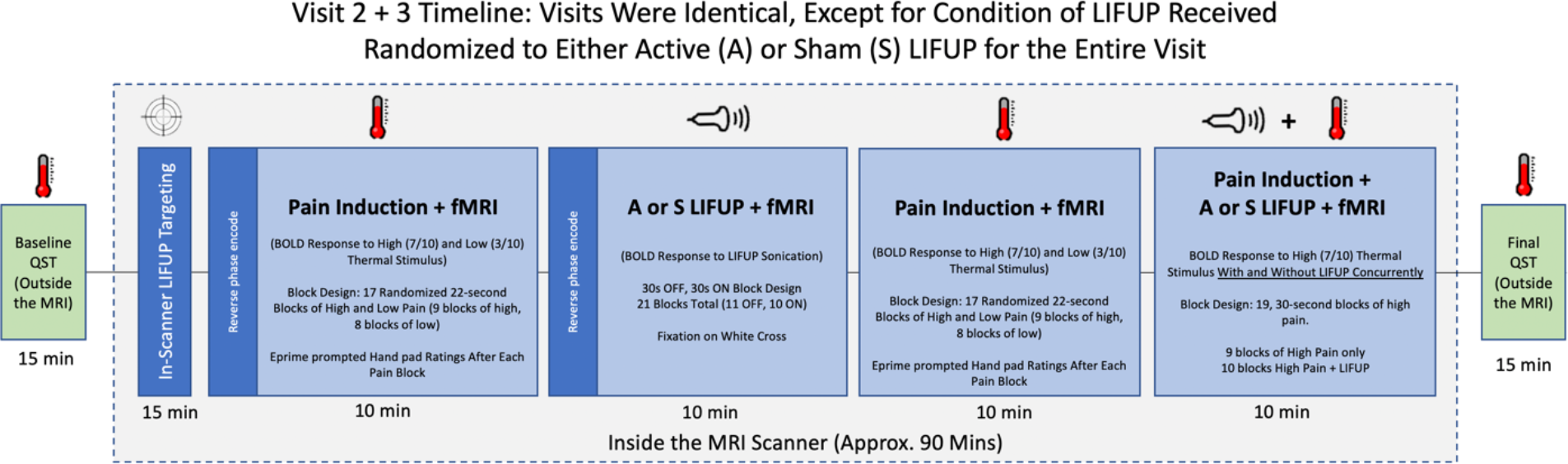
Study Overview. Experimental visits 2 and 3 were identical, with QST being conducted outside the scanner at baseline and after in-scanner LIFUP administration.

### Quantitative Sensory Thresholding (QST) – Outside of MRI Scanner

Quantitative sensory thresholding (QST) systematically determines information about Aβ fibers (sensory), Aδ fibers (pain), and C fibers (pain tolerance). We acquired threshold levels by attaching a 30x30mm thermode on the left forearm of participants (ATS thermode Medoc, Durham, NC). Using the Medoc Pathway System, we determined sensory, pain, and tolerance thresholds by administering incremental periods of thermal stimulus beginning at 32°C and increased by 0.5°C per second. Participants were asked to verbally indicate when the stimulus is detected (sensory), then when it is painful, and then to press a button when the stimulus was intolerable. When ‘intolerable’ was indicated, the thermode ceased heating and quickly returned to normal. This was repeated for 5 trials to obtain average sensory, pain, and tolerance thresholds. We repeated this at baseline and after LIFUP administration (approximately 90 minutes post-baseline QST, 45 minutes after initiation of first LIFUP sonication and 20 minutes after initiation of the second LIFUP session).

The 5 trial average at baseline was then used to run an additional program based on parametric estimation via sequential testing (PEST) to determine individualized pain ratings for each participant. We asked them to indicate at which heat level they perceived a 3 of 10 pain rating and a 7 of 10 pain rating. Studies have shown that stimuli rated as 7 out of 10 produce highly reproducible results in terms of pain ratings and fMRI activation without posing a significant risk to participants ^28^. These ratings were used in the concurrent pain/fMRI scanning protocol.

### Thermal Pain Administration during fMRI acquisition

For this study in the scanner we adapted an established fMRI thermal nociception model ^27^ as shown in **Figure 2a, b**. Cutaneous heat stimuli were applied via the Medoc Pathway System to the left forearm via a 30x30mm thermode. Pain levels were administered at individualized temperatures (low= 3/10, high =7/10) based on pre-determined ratings collected in the baseline QST paradigm.

**Figure 2.**
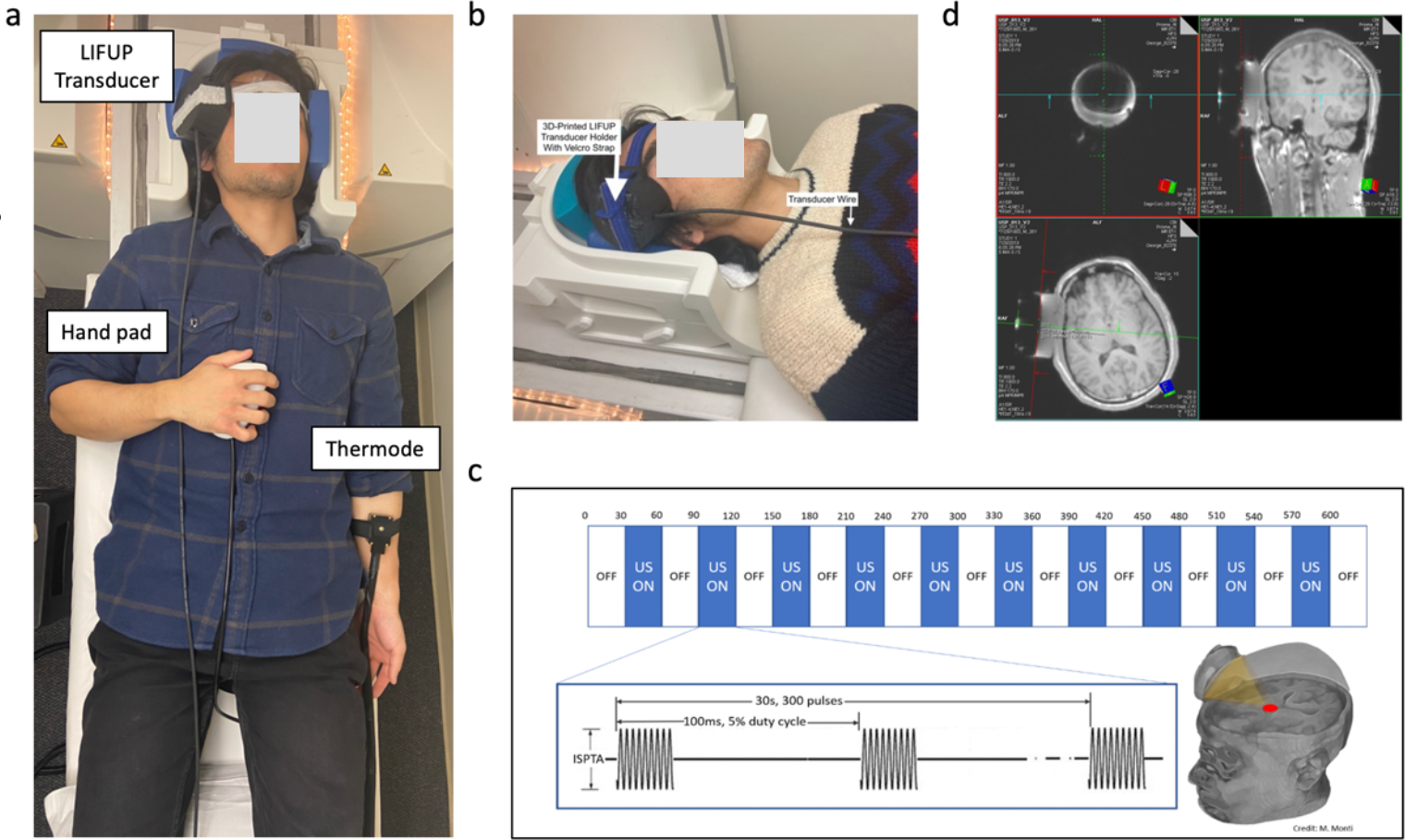
Administration of LIFUP in the MRI Scanner. a) compound picture of setup in scanner with thermode, hand pad, LIFUP device, b) Photograph of how LIFUP is attached to participant in the scanner head coil (32ch), c) Overview of LIFUP sonication block design paradigm, d) screenshot of the MRI console screen demonstrating how real-time LIFUP targeting is conducted.

For the first and third scan (“Pain Induction + fMRI, Figure 1), a total of 17 blocks of thermal stimuli were delivered, each lasting 22 seconds, while collecting fMRI data (TR: 1500 ms; TE 1: 15.20 ms, TE 2: 33.47 ms, TE 3: 51.74 ms; Voxel size: 3.0mm^3^; 38 slices, FA: 65 deg). Blocks were randomly assigned to be either low or high intensity, with low intensity corresponding to the participant’s “3 out of 10” and high intensity pain corresponding to “7 out of 10.” During these pain induction scans, a total of 10 high blocks and 7 low blocks were administered.

After each thermal stimuli, participants rated the intensity and unpleasantness of the pain from 1-5 using an MRI compatible hand pad in the right hand.

### Low Intensity Focused Ultrasound Pulsation (LIFUP)

#### LIFUP Targeting

An initial first MRI visit was conducted on all participants in order to acquire a structural scan (MPRAGE) to be used for outside of scanner targeting approximation. This involved using Brainsight (Rogue Company) to 3D reconstruct the structural scan and use a ultrasound transducer holder to mark the scalp target on the right temporal window that gives the nearest approximation to target the thalamus using an extrapolated straight line connecting the surface marker to the front 1/3 of the right thalamus. This scalp area was then marked with a permanent marker and used as the start reference for scanning. After marking, we connected the points using Brainsight and measured the distance from scalp to target, which determined whether a 1 or 2cm acoustic standoff pad would be needed. (As the puck focal length was fixed, we had to use this method to accommodate for different head sizes and shapes).

Inside the scanner, the transducer was coupled to the participant’s scalp using acoustic gel (Aquasonic) and an acoustic standoff pad in a custom 3D printed head-worn wearable mount that helps tune the distance of the ultrasound beam as the focal length was fixed (80mm) **(Figure 2c)**.

After the participant was in the bore of the scanner, a quick (1min) structural scout sequence (TR: 3.15ms; TE: 1.37ms; Voxel size: 1.6mm^3^; 128 slices, FA: 8 deg) was acquired and study personnel used the MRI Console computer to determine whether the transducer was correctly engaged with the pre-planned target. If there was deviation from the target, the study team would remove the subject from the scanner bore and while they were still on the gantry, readjust the head mount to better engage the target (anterior thalamus), repeat a scout image, and repeat this until they were certain the ultrasound was correctly positioned for that person.

#### LIFUP Administration

We administered ultrasound using the BrainSonix BXPulsar 1002 LIFUP System (BrainSonix Inc.) using a single-element, air-backed, spherical ultrasound transducer with a 61 mm diameter and 80 mm focal depth. The transducer was coupled to the scalp of the participant using a 3D-printed transducer holder that allows for sonication to occur in conjunction with ultrasonic standoff pads and ultrasound gel (**Figure 2b**).

Sonication parameters were as follow: [Fundamental frequency:650khz, pulse repetition frequency: 10 HZ, 10 HZ, Pulse Width: 5ms, Duty Cycle: 5%, Sonication Duration: 30s, Inter-Sonication Interval: 30 s, Number of Sonications: 10, ISPTA.3 719 mW/cm^2^]. LIFUP sessions were spread 10 minutes apart and were focused on the right anterior thalamus **(Figure 2d)**. E-Prime software was used to trigger LIFUP in a conventional 30s ON, 30s OFF block design.

#### Concurrent LIFUP/Pain/fMRI

The second 10-minute session of LIFUP (Pain Induction + LIFUP, **Figure 1**) was administered concurrently with a pain induction fMRI block design (TR: 1500 ms; TE 1: 15.20 ms, TE 2: 33.47 ms, TE 3: 51.74 ms; Voxel size: 3.0mm^3^; 38 slices, FA: 65deg). Participants during this concurrent multimodal scan received 19 total blocks of thermal stimulation, all at their self-reported “7 out of 10” intensity. During this pain induction, 10 of the pain blocks were administered concurrently with either active or sham LIFUP. Participants rated their pain intensity and unpleasantness on a hand pad rating from 1-5 after each block, irrespective of whether LIFUP was administered or not.

#### Blinding

Randomization was performed from MATLAB and entered into a spreadsheet and subjects were assigned numbers. One researcher (PS) knew the status of the subjects. However this researcher was in the control room behind the MRI scanner during the study and never interacted with the subject or the rest of the staff. During the scanning, PS either armed the BrainSonix, or not, depending on the subject’s status on that visit. He also checked before each study that the puck was working by placing the puck in a water bath and checking for waves.

### Statistical Analysis

The primary outcomes for this study was thermal sensory thresholds measured using the QST paradigm. These data were hand recorded then digitized for analysis using double data entry with spot checking to confirm accuracy.

Five consecutive trials of the thermal pain, sensory and tolerance threshold procedure were conducted at the beginning and end of each daily scanning session (Fig 1). The first of each of the 5 trials was discarded to preempt for novelty and orienting effects, as well as to ensure consistent A-delta fiber activation suppression during each testing period.

### Data Analysis

Linear mixed modeling was employed with unstructured covariance matrices to examine pre-post session effects, real versus sham stimulation effects and the pre-post X real-sham interaction while controlling for trial number. Participant intercepts, trial number and pre-post slopes were entered into the model as random effects at level-1.

## Results

### Demographics

29 subjects were consented. 19 completed the study and yielded usable data (11 women, mean age = 24.5 SD 4.6, range =18-38). The 10 who were not used in the final dataset were withdrawn for scheduling complications (n=6) or personal reasons (n=2) resulting in participant not completing one or both experimental visits after consent, scanner technical issues (n=1), or was withdrawn to a reported AE not directly caused by sonication (n=1). All of these participants were removed from the dataset before any analysis was conducted and before unblinding.

### Safety of concurrent LIFUP/MRI

There were no serious adverse events (AEs). One subject who was not included in the analysis experienced in-scanner panic attack sensations during an active LIFUP scan./ The participant experienced panic attack symptoms that began during non-LIFUP scans and culminated in his termination of scan. The subject’s symptoms resolved 15-minutes after the incident and was removed from the study.

Another subject contacted the study team to report a mild headache that persisted for 2 days following their sham LIFUP experimental visit.

### Location of the Sonication

Using a custom-written MATLAB-based graphical user interface, we analyzed the location of sonication for each individual to confirm thalamic targeting position. This method determines the angle of approach of the transducer and estimates the location of the peak sonication by calculating a beam 80mm from the center of the surface of the LIFUP transducer, along the path determined to be orthogonal to the transducer. The software then determines the central focal point of the area of sonication, which extends 1.5cm at a width of 0.5cm **(Figure 3a)**.The program then gives, as output, the location of stimulation (in pixels and mm), distance from stimulation site to target (anterior thalamus), and overlap between the area of stimulation and the anterior thalamus (in raw area, as a percentage of total area of the anterior thalamus, and as a percentage of the stimulation area that falls within the target). The target engagement data is then overlaid on a standard brain, revealing the projected sonication beam reliably engaged the anterior thalamus **(Figure 3b)**.

**Figure 3.**
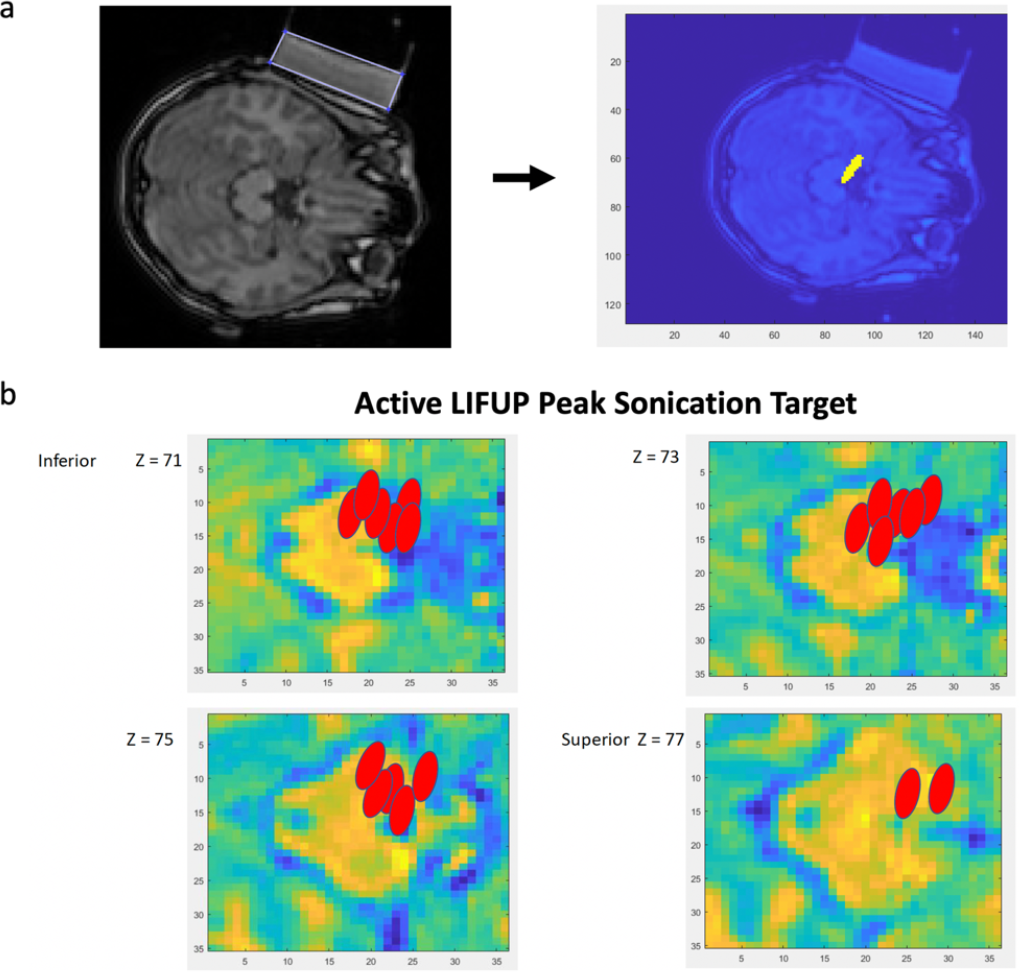
Software-based confirmation of deep brain sonication. a) individual MRI images were used to create trajectory models of the sonication beam and b) the center of the sonication beam coordinates were then mapped onto a standard brain.

### Quantitative Sensory Thresholds (QST)

We used linear mixed modeling with unstructured covariance matrices to examine pre-post session effects, real versus sham stimulation effects and the pre-post X real-sham interaction while controlling for trial number. Participant intercepts, trial number and pre-post slopes were entered into the model as random effects at level-1.

For thermal sensory thresholds (“indicate when can you feel the initiation of thermal stimulus”), while controlling for trial number (F(1,14.16)=32.97, p<.0001), no main effects for pre-post stimulation (F(18.08)=1.79, ns) nor real versus sham were found (F(1,250.55=1.98, ns). The pre-post X real-sham interaction trended toward significance (F(1,250.54=3.55, p=.06). In the real stimulation condition, sensory thresholds increased, on average 0.83°C (SE=.40) after the session, whereas, in the sham group, sensory thresholds increased 0.12°C (SE=.38). These sensory changes were in the hypothesized direction but did not meet significance.

With respect to thermal pain thresholds (“indicate when you find the stimulus painful”), a main effect for pre-post was observed (F(1,17.76)=8.97, p=.008) but not real-sham (F(1,245.95)=0.50, ns) while controlling for trial number (F(1,12.60)=24.21, p<.0001). Moreover, the pre-post X real-sham interaction was significant (F(1,245.95)=4.03, p=.046). **(Figure 4)**

**Figure 4.**
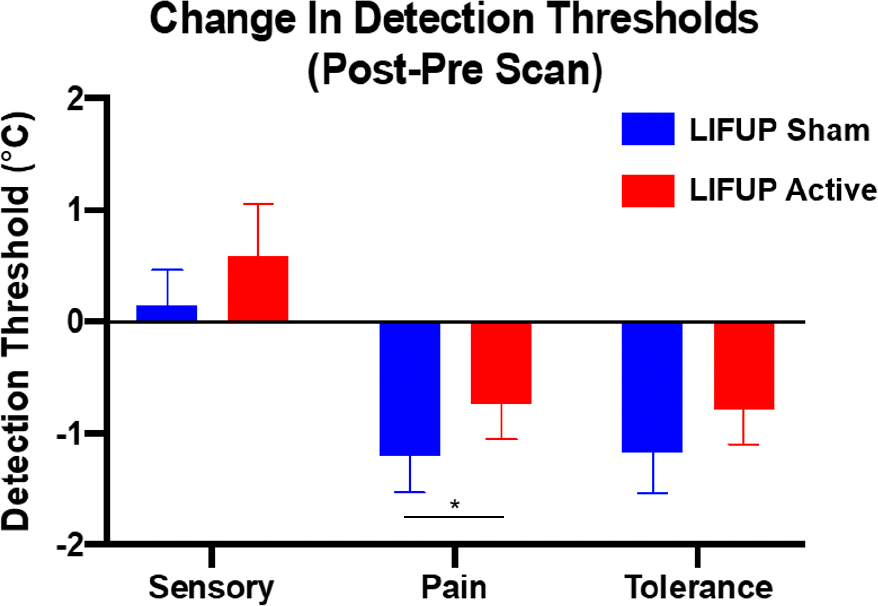
Overall change in QST pain threshold by condition. Temperature sensitivity increases were significantly attenuated (timeXcondition p=0.046) after active LIFUP (0.51 degree change) relative to sham stimulation (1.08 degrees).

The whole 90-minute procedure caused subjects to need less heat after the complete study to register it as painful. In the sham stimulation condition, thermal pain thresholds decreased 1.08°C (SE=.28) pre-post session, but only decreased .51°C (SE=.30) pre-post session in the real stimulation group. That is, LIFUP significantly attenuated the increase in sensitivity that occurred as a result of the 2-hour study procedures.

With respect to thermal pain tolerance (“indicate when you can no longer stand this pain, please stop now”), there was a main effect for pre-post session on thermal pain tolerance (F(1,17.93)=6.51, p=.02) but not real-sham (F(1,247.51)=2.05, ns) while controlling for trial number (F(1,443.23)=33.22, p<.0001). The real-sham X pre-post interaction was not significant (F(1,247.46)=1.15, ns). Mathematically, on the day they received LIFUP, subjects tolerated a higher temperature after the stimulation than on the day they received sham. The sham stimulation condition was associated with a decreased pain tolerance of 0.85°C (SE=.24) pre-post session, while real stimulation was associated with a decrease of 0.60°C (SE=.31) pre-post session (NS).

In summary, there was a significant antinociceptive effect of LIFUP on pain thresholds. However LIFUP did not significantly change sensory or tolerance thresholds, although the group differences for tolerance were in the direction of an anti-nociceptive effect.

Handpad Ratings: There was no significant effect of LIFUP on hand pad ratings conducted within the scanner (1-5 VAS rating scale).

### Integrity of the Blind

Subjects were asked after each MRI LIFUP visit whether they thought they received active or sham, and how confident they were on a scale of 1-10. All staff except PS were kept blind to randomization until all data were entered and checked and discrepancies resolved.

<u>Best guess</u> – 6 subjects declined to guess which intervention they received after the first visit, and 2 declined after the second visit. Overall, subjects were not confident in their guesses (average confidence score 4.13/10). Despite the low confidence, overall and as a group, subjects were able to guess better than chance which intervention they received. Combining guesses from both visits, more subjects assigned sham thought they received sham (32%), and more subjects assigned real thought they received real (42%) than should have occurred by chance (chi-square 7.8, 3DF, p=0.04). In examining the comments about what informed their guess, two issues emerged. Some subjects thought they heard a clicking or humming sound, which they presumed was active. Notably one subject guessed wrongly that they had received real based on this noise when in fact they received sham. More interestingly, of the 5 sessions where subjects rated a 9 or 10 confidence, they based their decision on the perception that the pain felt less during that run, and they guessed real.

## Discussion

This study was an initial test of LIFUP’s ability to stimulate a deep nucleus and produce observable behavioral changes in healthy adults. We found that two, 10 minute sessions of LIFUP delivered safely to the right anterior thalamus, produced significant antinociceptive effects in pain threshold ratings measured 25-45 minutes after sonication. MRI-guided LIFUP administered within the guidelines of FDA power intensity is thus feasible, and safe, with no serious adverse events related to LIFUP.

Although we found an effect on pain threshold over 90 minutes and the two LIFUP sessions, we did not find an immediate effect of LIFUP concurrent with pain. The antinociceptive effects were not immediate but rather delayed and likely cumulative. This is consistent with data from frog sciatic nerve where LIFUP causes a reduction in excitability that persists for up to 45 minutes.^29^

### Limitations

There are several limitations that should be kept in mind when considering this study and our conclusion that LIFUP of the anterior thalamus can reduce pain.

Although we performed this study inside the MRI scanner, the targeting could be improved in the future. It appears that most subjects had the LIFUP focus within the thalamus. We did not perform thermal imaging to confirm our location. Additionally, we did not stimulate another brain region as a control, so theoretically this could be a global LIFUP effect and not due to thalamic stimulation. Studies with active brain regions as control conditions are needed for better regional brain behavior causal statements.

This was a relatively small sample size and we were only powered to detect large effects. Additionally, this was a limited dose of LIFUP, bound in part by safety considerations in this new field. As more studies are done and the safety database grows, higher doses can be delivered, which may have larger effects.

Our temporal understanding of the LIFUP induced changes is limited. Because this was done in the scanner, no QST was delivered between the two doses. Thus, we do not know if these two doses were additive or not, or how long it takes for the antinociceptive effects to occur. Similarly, we did not perform later tests that day or the following day to test how long the effect might last. A follow-up study in the pain lab outside of the scanner would allow one to administer the QST more frequently and obtain finder grained temporal understanding of the effects.

The MRI scanner is a loud and noisy environment, and subjects wore earplugs. Some have reported that certain parameters of LIFUP can produce noise carried via bone conduction. Future studies will need to potentially control for this confound, perhaps employing active noise masking via headphones.

We did not find a large and immediate effect like is sometimes seen with thalamic ablation for pain. It is likely that we are not fully blocking all pain signals, and are instead modulating only a subset.

Further follow-up studies are now needed, with refined targeting, some performed out of scanner, with dose finding.

## Conclusions

Two 10-minute sessions of LIFUP delivered to the anterior thalamus significantly changes pain thresholds in healthy volunteers. LIFUP appears to be a promising new form of brain stimulation able to focally target deep brain structures. Further work needs to explore dosing and parameter optimization in order to move LIFUP forward as a potential therapy for pain disorders.

## Data Availability

The data that support the findings of this study are available from the corresponding author, BWB, upon reasonable request.

